# Evidence of early community transmission of Omicron (B1.1.529) in Delhi- A city with very high seropositivity and past-exposure!

**DOI:** 10.1101/2022.01.10.22269041

**Authors:** Rahul Garg, Pramod Gautam, Varun Suroliya, Reshu Agarwal, Arjun Bhugra, Urvinder S. Kaur, Santanu Das, Chhagan Bihari, Anil Agarwal, S. K Sarin, Ekta Gupta

## Abstract

**Background:** Since identification, infections by new SARS-CoV-2 variant Omicron are rapidly increasing worldwide. There is huge gap of knowledge regarding virus behaviour in the population from low and middle income countries. Delhi being unique population with a high seropositivity and vaccination rate against COVID-19 infection. We aimed to study the epidemiological and clinical presentations of few early cases of community spread of Omicron infection in the state.

**Methods:** This is a prospective study where respiratory specimen from all RT-PCR confirmed positive cases between November 25^th^-December 23^rd^ 2021 collected from five districts of Delhi were subjected to whole genome sequencing. Complete demographic and clinical details were recorded. We also analyzed the formation of local and familial clusters and eventual community transmission.

**Findings:** Out of the 264 cases included during study period, 68.9% (n=182)were identified as Delta and its sub-lineages while 31.06% (n=82) were Omicron with BA.1 as the predominant sub-lineage (73.1%). Most of the Omicron cases were asymptomatic (n=50,61%) and not requiring any hospitalizations. A total of 72 (87.8%) cases were fully vaccinated. 39.1% (n=32) had a history of travel and/or contacts while 60.9 (n=50) showed a community transmission. A steep increase in the daily progression of Omicron cases with its preponderance in the community was observed from 1.8% to 54%.

**Interpretation:** This study is among the first from India to provide the evidence of community transmission of Omicron with significantly increased breakthrough infections, decreased hospitalization rates, and lower rate of symptomatic infections among individuals with high seropositivity against SARS-CoV-2 infections.

## 1. Introduction

Since the onset of the COVID-19 (Coronavirus Disease) pandemic in December 2019, many variants of Severe Acute Respiratory Syndrome Coronavirus-2 (SARS-CoV-2) have emerged. On December 26^th^2021, approximately two years since the first reported case and after a global estimated 260 million cases resulting in 5.2 million deaths, World Health Organization (WHO) reported a new SARS-CoV-2 variant of concern (VoC), Omicron^1^. Omicron emerged in a COVID-19-weary world, rife with frustration and anger with overall negative impacts on social, mental, and economic wellbeing.

The first sequenced omicron case was reported from Botswana on November 11^th^2021, followed by another case reported from Hong Kong in a traveller from South Africa^2^. Approximately one week later, on December 2^nd^2021, the first two Omicron cases in India were identified in the state of Karnataka^3^. On December 5^th^2021, New Delhi identified national capital’s first and India’s fifth case of Omicron from a traveller returning from Tanzania^4^. By December 30^th^, India had recorded 961 cases across 21 states and union territories, out of which 263 cases were registered in New Delhi^5^.

New Delhi being unique with a seropositivity rate of 89.5% and having gone through a colossal and deadly outbreak of Delta infections in the months of April-June 2021, community spread of omicron could be interesting and different from other places^6^. Therefore, in the present report we highlight the epidemiological and clinical presentation of few early cases of community acquired infections of Omicron in the Delhi State.

## 2. Methodology

The Department of Clinical Virology, at the Institute of Liver and Biliary Sciences, New Delhi (ILBS) is an ICMR (Indian Council of Medical Research) designated COVID-19 diagnostic facility and a part of National consortium, INSACOG (Indian SARS-CoV-2 Genomics Consortium). Respiratory specimen from RT-PCR confirmed cases (November 25^th^ to December 23^rd^ 2021) which were being referred to our laboratory from designated districts of Delhi (South, South East, South West, East and West) as per the Government of National Capital Territory of Delhi (GNTCD) directives, were subjected to whole genome sequencing (WGS). These cases do not include those international travellers that tested positive at the time of entry at the airport, however if they turned out to be positive in due course in the follow-up screening were included in the study. This time frame (November 25^th^ to December 23^rd^ 2021) was chosen keeping in mind that it represents the period during which an Omicron variant was first designated as VoC by WHO and the last date of specimen collection used to generate genome sequence data for this manuscript.

The complete demographic and clinical details of patients infected with SARS CoV-2 were recorded from Specimen Referral Form (SRF) issued by ICMR (Indian Council of Medical Research, New Delhi) and the patient information sheet (PIF) sent from each testing site along with samples. Any missing data from the form of all the enrolled cases were recorded via telephonic interviews.

### 2.1 Nucleic acid extraction

Total viral RNA was extracted using automated nucleic acid extraction platform Chemagic Viral DNA/RNA kit (Perkin Elmer, Waltham, MA, USA) on a Chemagic 360 instrument (Perkin Elmer, Waltham, MA, USA) following manufacturer’s instructions maintaining adequate bio safety precautions.

### 2.2 SARS-CoV-2 Whole Genome Sequencing

Sequencing of the viral isolates was done by Illumina COVIDSeq protocol on NextSeq 550 platform using ARTIC V3 Primers as per the manufacturer’s instructions. The quality check of the prepared libraries was performed using DNA high sensitivity assay kit on Bioanalyzer 2100 (Agilent Technologies, United States). The concentration of the libraries was assessed on Qubit (Thermo Fisher Scientific Inc., USA).

#### 2.2.1 Quality control, Mapping of sequences and Lineage assignment

The raw data in the form of binary base call format (.bcl files) was generated from the NextSeq 550 instrument. These raw files were converted, demultiplexed to fastq file using bcl2fastq (Illumina, v2.20) and were aligned against the SARS-CoV-2 reference genome (NC_045512.2). The alignment of unmapped reads to a reference genome and generation of a consensus genome sequence was done within the custom Illumina BaseSpace Sequence Hub. The lineages nomenclature for each sequence was done using latest pangoLEARN version as and when available using either local installation of stand-alone pangolin workflows or Illumina® DRAGEN COVID Lineage App (v3.5.5) following the default parameters.In case of DRAGEN COVID Lineage tool, the minimum accepted alignment score was set to 22 and results with scores <22 were discarded. The coverage threshold and consensus sequence generation threshold were set to 20 and 90 respectively. The lineage calling target coverage which specifies the maximum number of reads with a start position overlapping any given position was set at 50.

Samples with a minimum of 30x genomic coverage in > 50% of sequence and % of non-N bases (Coverage >= 10x) > 80% were finally selected for downstream analysis. The median genomic coverage of quality passed samples was 98.8 while the median sequencing depth was 1805. The retrieved sequences were deposited in the public repository, GISAID (https://www.gisaid.org).

#### 2.2.2 Construction of phylogenetic tree

Using consensus FASTA generated from BaseSpace pipeline, the phylogenetic tree was build following the Nextstrain protcol for genetic epidemiology of SARS-CoV-2 using the reference genome of SARS-CoV-2 isolate named Wuhan-Hu-1 (NCBI Reference Sequence: NC_045512.2) (8). The tree was visualised in Auspice.

### 2.3 Mapping of cases by geographical location

The identified Omicron cases were geotagged using the address available in the metadata. Before tagging the location, all the personal details of cases were removed. Only the travel/contact history as revealed by the subject in the SRF form was used. The family relationship of each sample was deduced from having the same or different address. A local cluster was defined by having three or more Omicron cases in the close neighbourhood.

### 2.4 Ethical approval

The present study was approved by Institutional Ethics Committee (IEC) at Institute of Liver and Biliary Sciences, New Delhi (Ethical no. IEC/2020/77/MA07). In this study de-identified and anonymised samples were subjected to whole genome sequencing.

### 2.5 Statistical analysis

Data recorded in the PIF was digitized for analysis using SPSS version 15 (IBM, Bangalore, South Asia). Descriptive statistical tools were used to estimate the frequencies of categorical variables. Median and interquartile ranges (IQR) were reported for continuous variables. Chi-square test and univariate analysis were used to measure the association between the study variable and categorical outcomes. A p-value of <0.05 was considered statistically significant.

## 3. Results

### 3.1 Baseline characteristics of studied cases

A total of 332 samples were referred for WGS from November 25^th^ till December 23^rd^, 2021 from different testing laboratories across the five districts of Delhi. Out of them, 264 quality checked passed samples were included for the analysis. Baseline characteristics of the studied population are described in Table 1, which clearly indicate that 68.9% (n=182) adult population (aged 18-60 years) was more infected in comparison to paediatric and elder population. A total of 80.6% (n=231) of the infected cases were fully vaccinated and 12.9% (n=20) were reported to have previous SARS-CoV-2 infection. Overall 58.7% (n=91) cases were symptomatic with 9.6% (n=15) requiring hospitalization, majority of them with underlying comorbidities. Mortality was noted in 1.2% (n=2) of the cases, infected with Delta variant.

**Table 1.**
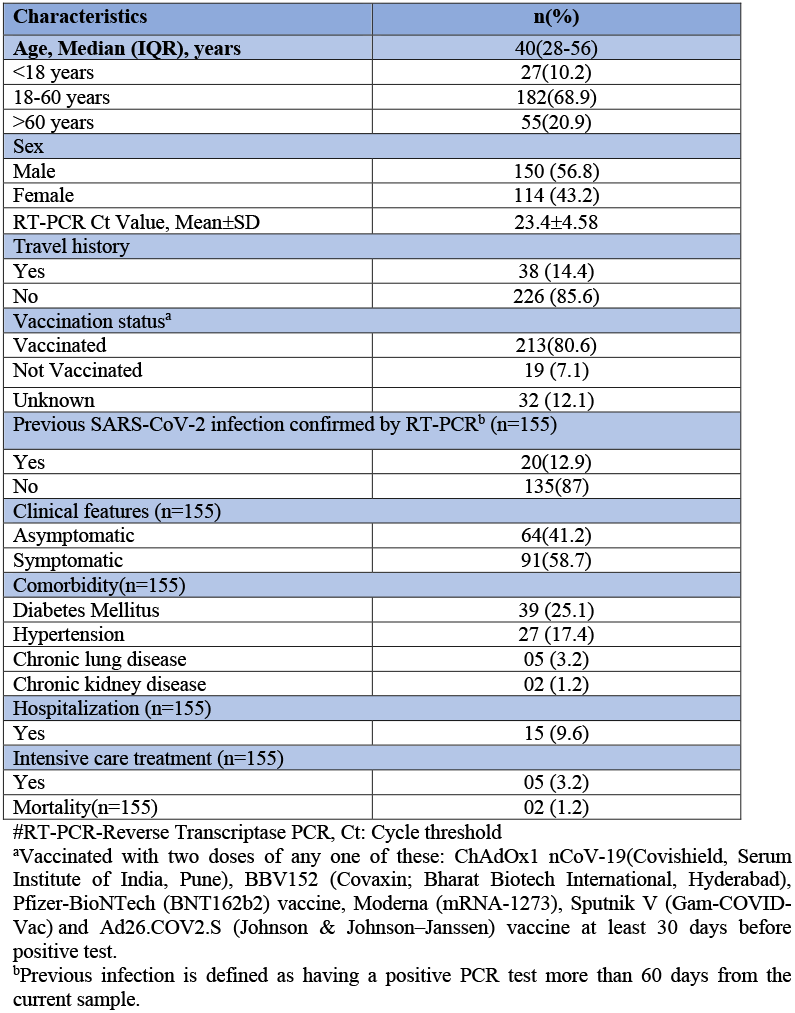
Baseline characteristics of SARS-CoV-2 infected cases

### 3.2 Distribution of varied lineages of SARS-CoV-2 across five districts

Out of 264 samples sequenced, 68.9% (n=182) were Delta and its sub-lineages B.1.617.2 while rest 31.06% (n=82) were Omicron (B.1.1.529) variant (Figure 1). Out of the total Omicron cases, 73.1% (n=60) were of BA.1 (Next stain clade 21K) while 26.8% (n=22) were of BA.2 lineage (Next stain 21L) (Figure 2).

**Figure 1:**
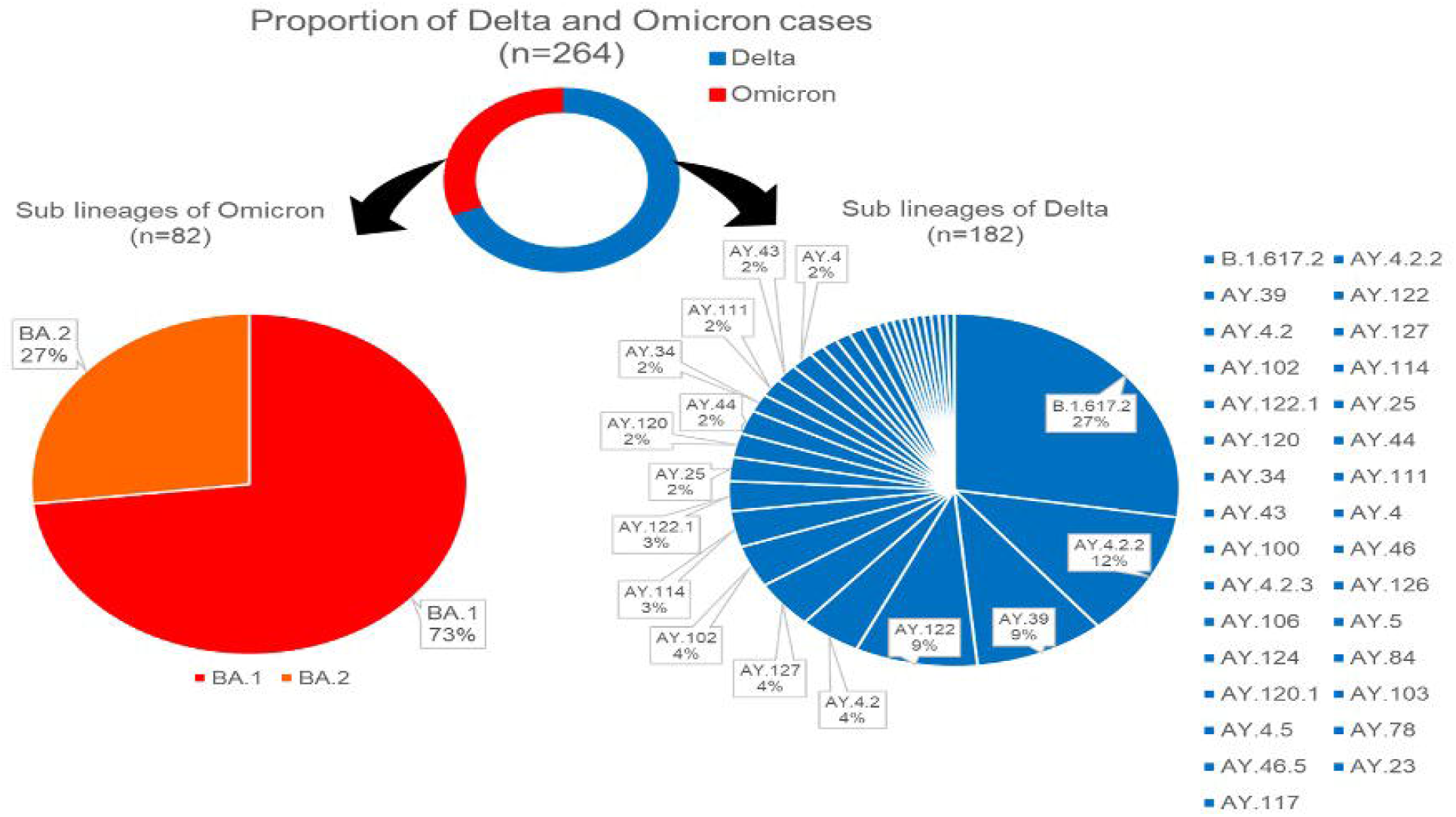
Variant distribution with lineages and sub-lineages of 264 cases

**Figure 2:**
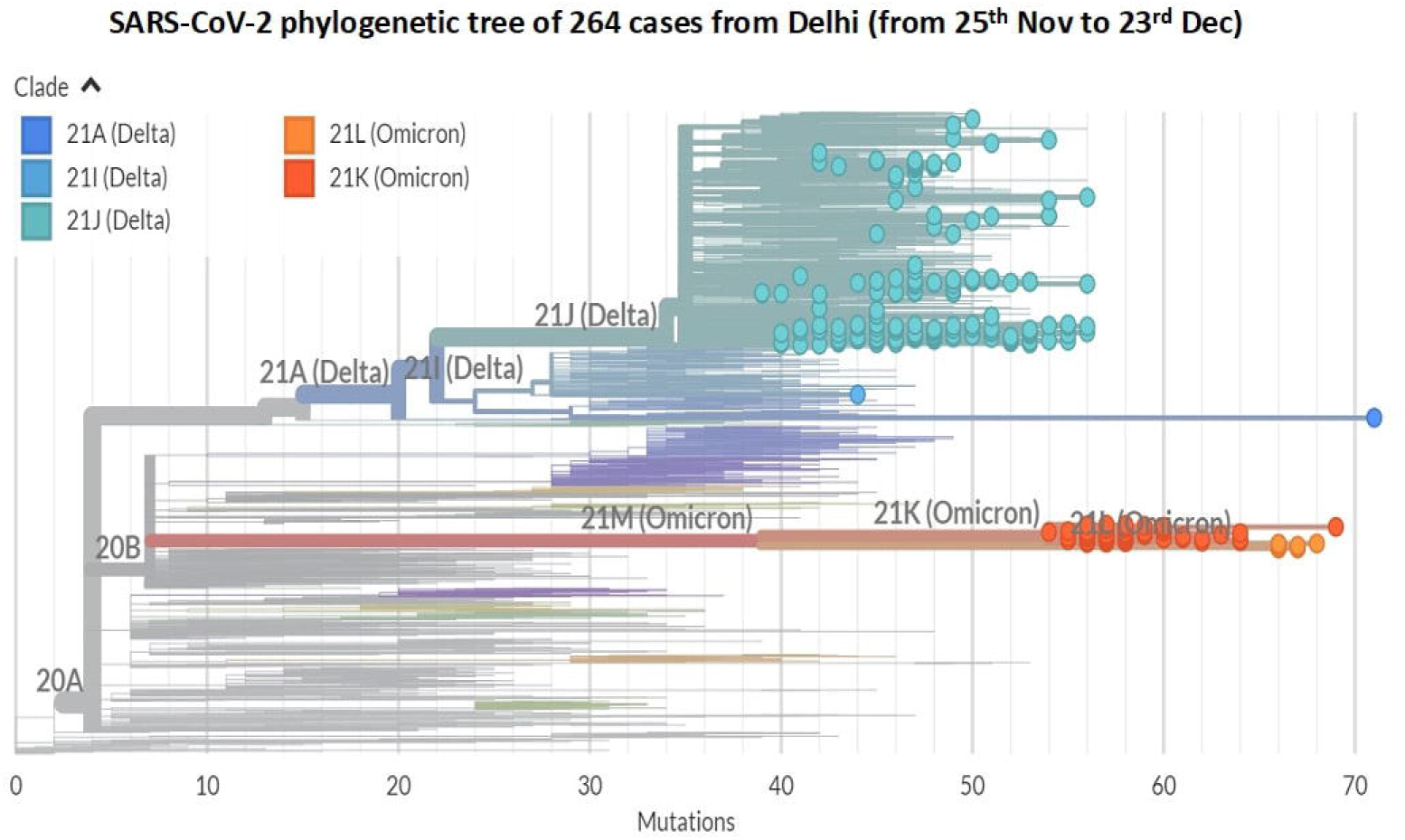
Phylogenetic representation of all the cases in the study.

Majority of cases belonged to South (45%) followed by West (26%) district of Delhi (Figure 3A). The detailed description of detected lineages across the five districts had been shown in Figure 3B.

**Figure 3.**
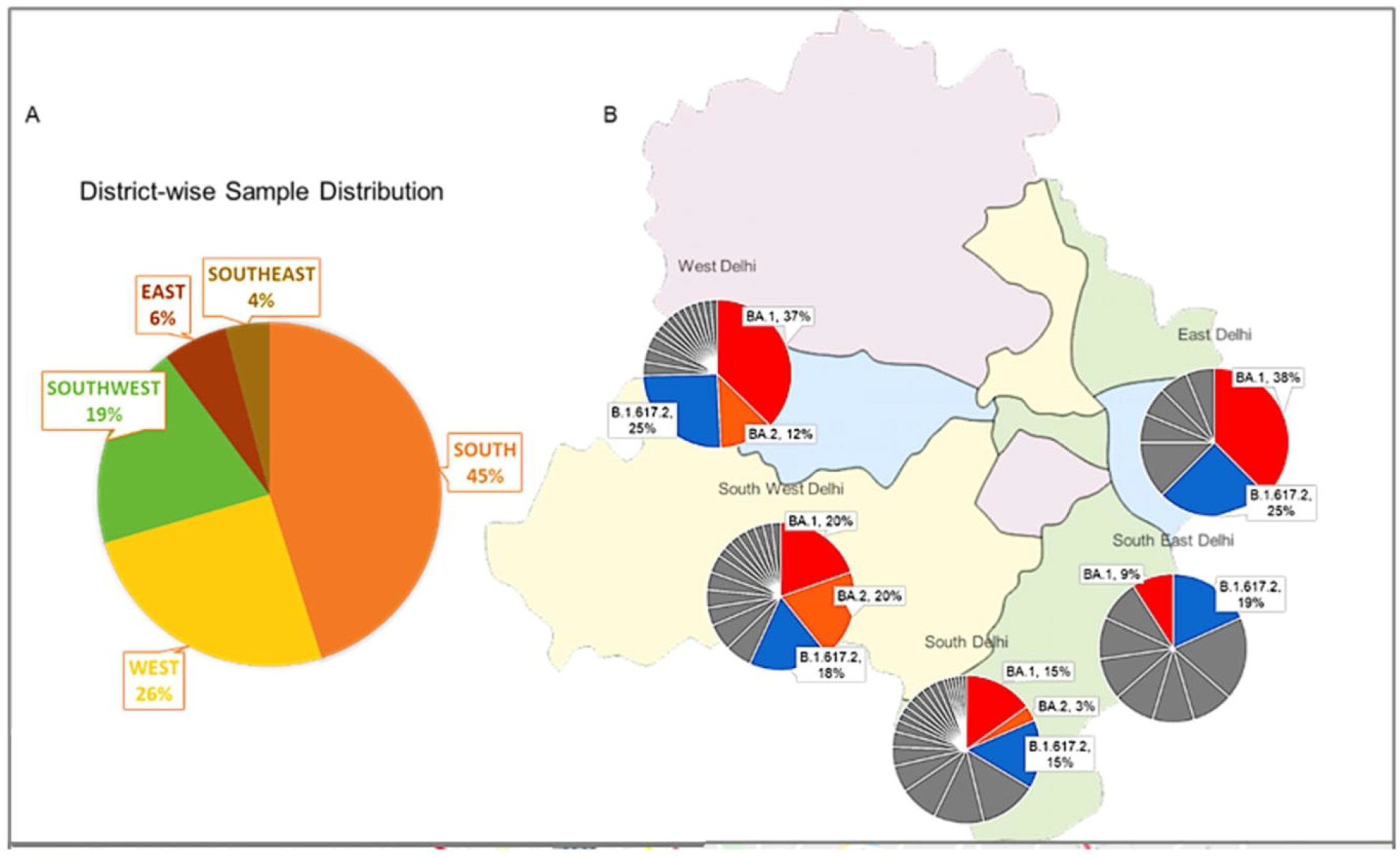
Distribution of SARS-CoV-2 lineages and sub-lineages among five studied districts. 3A. The district-wise distribution of cases (n=264). 3B. The sub-lineage distribution in five districts of Delhi. The Omicron BA.1 is shown in red and BA.2 in orange colour. The Delta parent lineage (B.1.617.2) is shown in blue and rest Delta sub-lineages in grey colour.

### 3.3 Clinical characteristics of Omicron cases

The median age of Omicron infected cases was 35 years (IQR 24-53) with male (57.3%, n=47) predominance. Demographic description of all 82 Omicron cases is depicted in Table 2. Majority of the cases (n=50, 61%) were asymptomatic. Hospitalization was recorded in only 3.6% (n=3) cases with diabetes mellitus and hypertension as an underlying comorbidity. None of the admitted cases required intensive care throughout their hospital stay. Time since RT-PCR negativity was 10±3days. Looking at the vaccination status, total of 87.8% (n=72) cases were fully vaccinated and an additional 01 (1.2%) had received heterologous vaccination (2 doses of mRNA-1273 [Moderna] and 1 dose of BBV152 Covaxin vaccine).

**Table 2.** Baseline characteristics of Omicron cases.

| Characteristics | N(%) |
| --- | --- |
| **Age, Median (IQR), years** | 35(24-53) |
| <18 | 12 (14.6) |
| 18-60 | 56 (68.3) |
| >60 | 14 (17.1) |
| **Sex** |  |
| Male | 47 (57.3) |
| Female | 35 (42.7) |
| RT-PCR Ct Value, Mean±SD | 23.9±4.1 |
| **Vaccination status** |  |
| Vaccinated | 72 (87.8) |
| Not Vaccinated | 10 (12.2) |
| **Travel history** |  |
| Yes | 32 (39.1) |
| No | 50 (60.9) |
| **Previous SARS-CoV-2 infection confirmed by RT-PCR** |  |
| Yes | 14 (17.1) |
| No | 68 (83.9) |
| **Clinical features** |  |
| Asymptomatic | 50 (61.0) |
| Symptomatic | 32 (39.0) |
| **Comorbidity\*** |  |
| Comorbidities | 15 (18.3) |
| No comorbidities | 67 (81.7) |
| Hospitalization | 03(3.6) |

RT-PCR-Reverse Transcriptase PCR, Ct: Cycle threshold
\*Co-morbidity was defined as $\geq 1$ of the following conditions: hypertension, diabetes, chronic lung disease, chronic cardiac disease, chronic kidney disease.

### 3.4 Epidemiological description of Omicron cases

In the present study, the first two cases of Omicron were detected during the first week of December with travelling history from South Africa. Afterwards we observed a steep increase in the daily progression of Omicron cases with its preponderance in the community from 1.8% to 54% (Figure 4).

**Figure 4.**
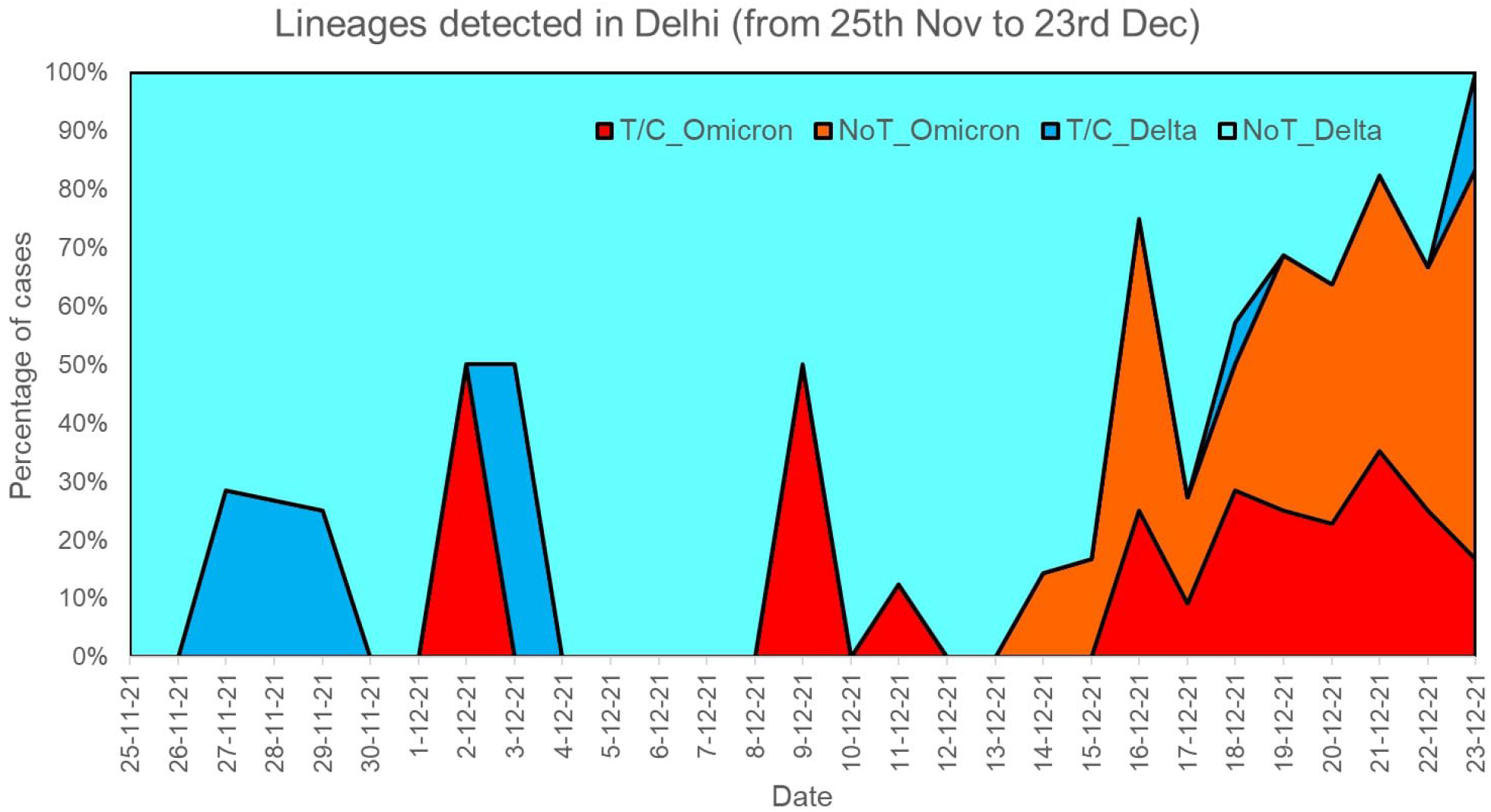
Daily progression of SARS-CoV-2 lineages with respect to travel history and community transmission. T/C-History of Travel and/or Contact; NoT-No history of travel and/or contact

History of international travel was documented among 19 cases [South Africa (n=6),UK (n=5), UAE (n=4), USA(n=3) and Canada(n=1)] while thirteen cases were the contacts of such travellers. On comparing cases with history of recent international travel and/or their contacts (n=32) with no travel history (n=50), no statistically significant difference could be ascertained (Table 3). However, the proportion of asymptomatic cases (n=23, 71.8%) was higher among travellers/ and or contacts in comparison to the community (n=27, 54%), suggesting acquisition of new variant by some cases from abroad with local importation.

**Table 3.**
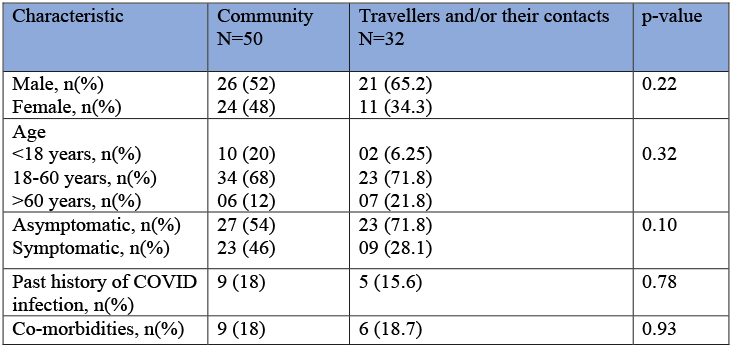
Epidemiological characteristics of cases with travel history and/ or contacts and with no travel history (community).

Among total of 82 Omicron cases, 87.8% (n=72) were fully vaccinated. ChAdOx1 nCoV-19 (Covishield) was the most common administered vaccine in around 56% of cases. The other vaccines included BBV152 Covaxin (12%), BNT162b2 [Pfizer] (11%), mRNA-1273 (Moderna) (4%), Sputnik V vaccine (Gam-COVID-Vac) (4%) and Ad26.COV2.S vaccine (Johnson & Johnson–Janssen) (1%).

### 3.5 Distribution of Omicron cases across five districts of Delhi

On describing the geographical location of the Omicron cases, they were identified widespread across the state, but most local clusters (≥ 3 cases in close vicinity from unrelated family) were from West (40.2%) and South (26.8%) districts of the Capital (Figure 5).

**Figure 5:**
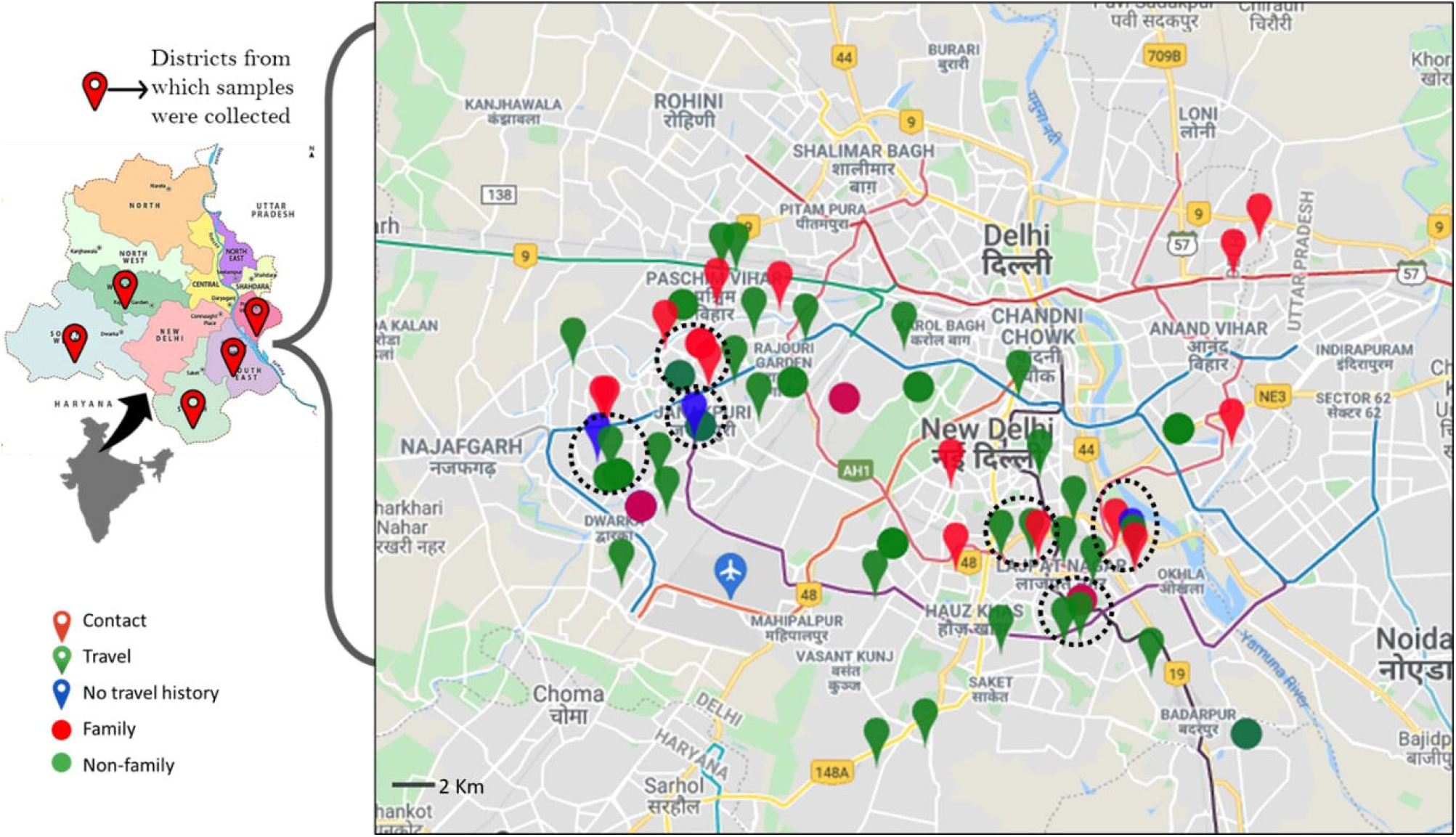
Geotagging of Omicron cases (n=82) from five districts of National Capital, Delhi. Black dotted circles represent local cluster with three or more cases in close vicinity. Red solid circles denote multiple cases from same families. Cases are labeled based on confirmed travel history (Green), Contacts of cases with travel history (Red) and cases with no travel history (Blue).

### 3.6 Family cluster of Omicron cases

Among a total of 82 Omicron cases, 46.3% (n=38) belonged to a total of 14 families (Average member/family = 2.7). Out of these 14 family clusters, only 4 families (5 individuals) had documented travel history from different countries: 1 member from United Kingdom, United States of America, United Arab Emirates each and 2 members of same family from South Africa. Out of the remaining 10 families with no international travel history, three families contracted Omicron following contact with a non-family member with international travel history. Rest of the 20 individuals from 7 families contracted Omicron infection possible due to community transmission (Figure 6).

**Figure 6:**
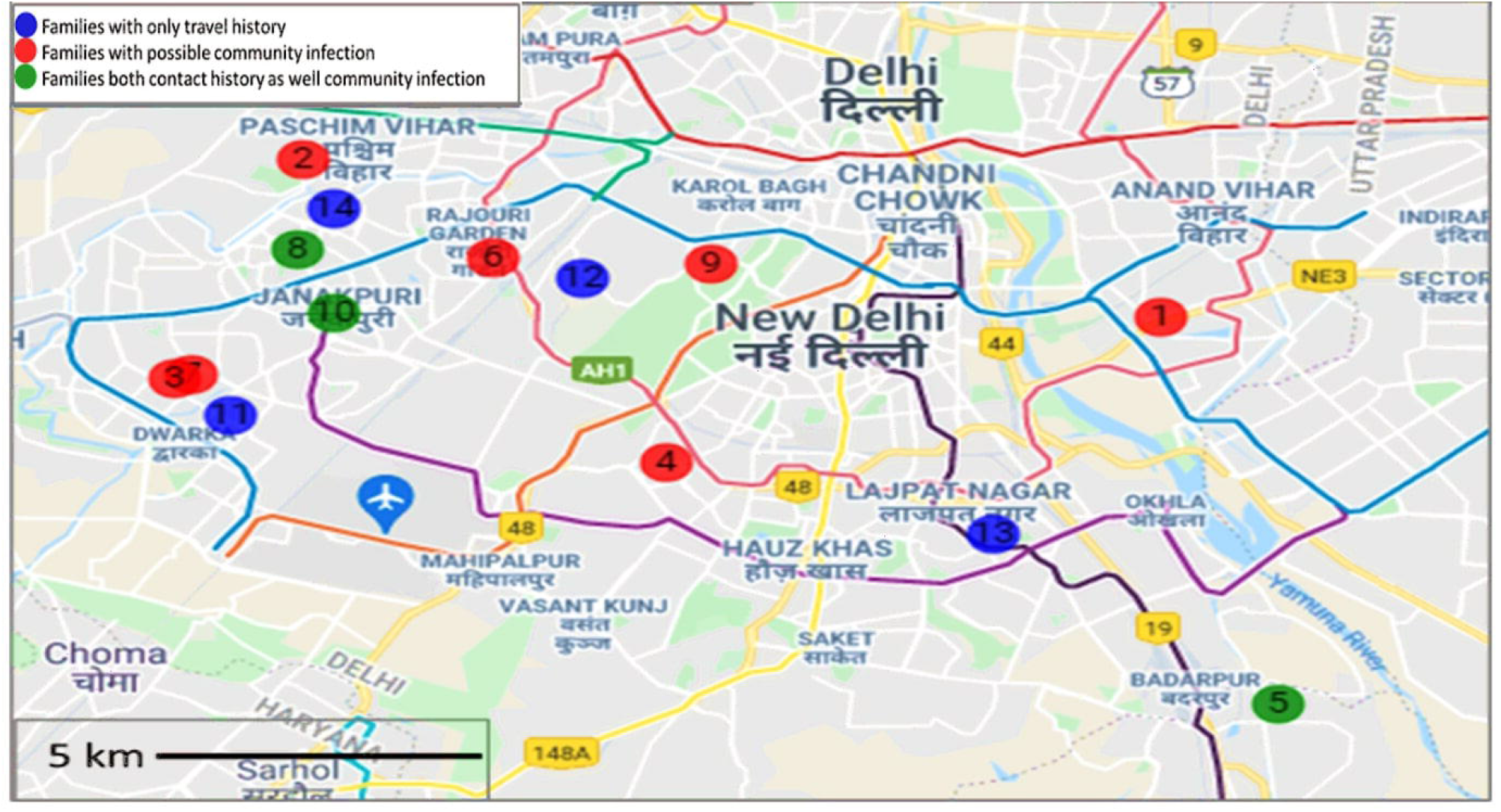
Geotagging of family clusters in Delhi. A total of 14 families with 38 Omicron cases were identified. Plotted here are families colored by their respective travel history, possible community acquired infection and both

## Discussion

Since the first confirmed Omicron reported from Delhi, a steep increase in the number of cases has been observed (Figure 7). To the best of our knowledge this is the first study from India to provide the evidence of community transmission of Omicron with significantly increased breakthrough infections, decreased hospitalization rates, and lower rates of symptomatic infections.

**Figure 7:**
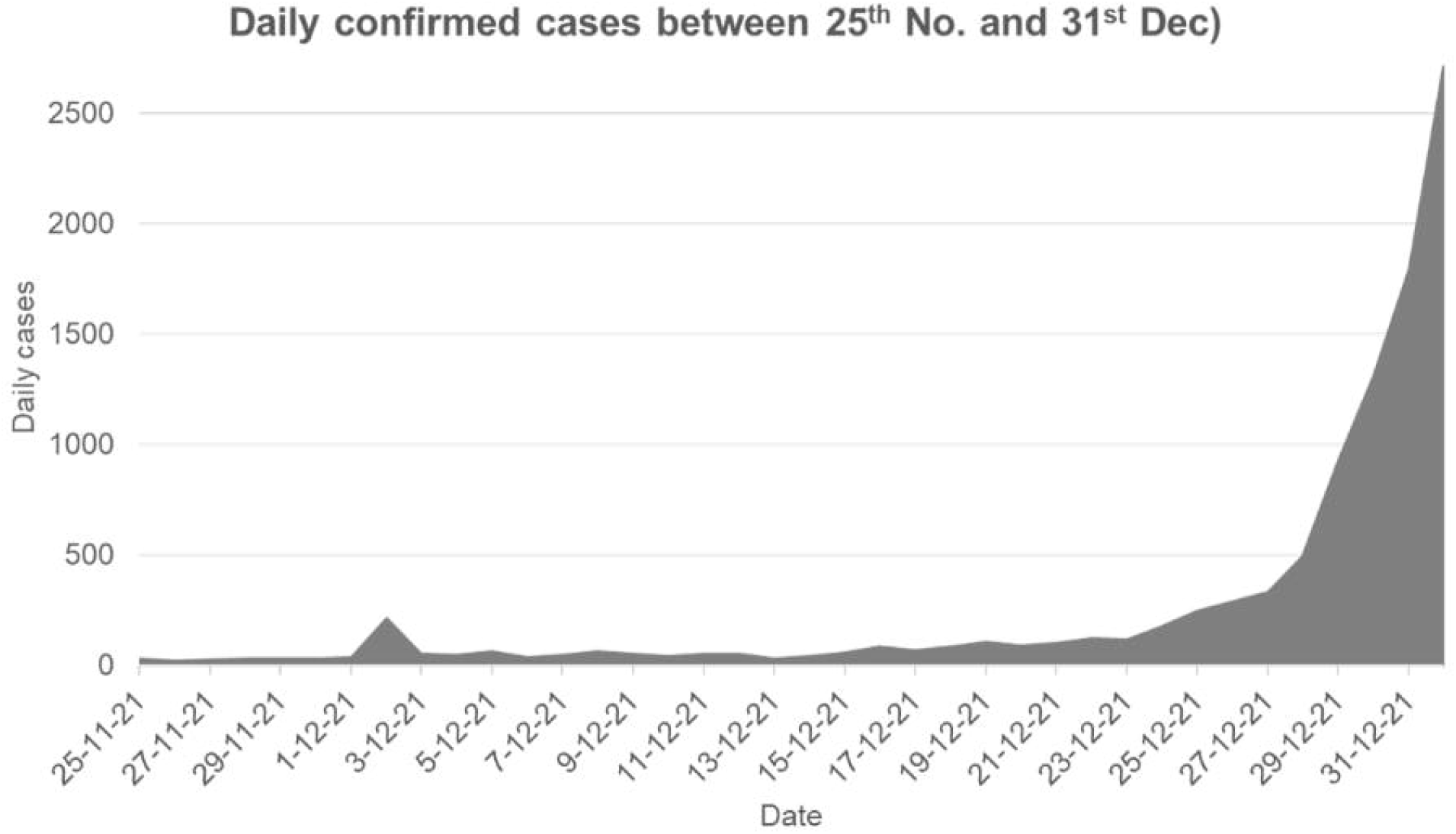
Progression of daily recorded confirmed Covid-19 cases in Delhi from November 25^th^– December 31^st^ 2021[Referred from https://covid19bharat.org/state/DL]

On December 5^th^ 2021, the national capital New Delhi identified the state’s first and country’s fifth case of Omicron from a traveller returning from Tanzania^4^. Since then the cases are increasing with a doubling growth rate of 3-4 days, and replacing Delta as the dominant VoC in the time span of less than a month. Our study is based on epidemiological, clinical and genome sequence analysis of 264 cases since the day Omicron was designated as VoC by WHO.

We observed a definite shift from Delta predominance^7^ to Omicron along with its community spread. Similar findings were observed from various part of the world including South Africa and Denmark where the new variant simultaneously emerged at the end of November 2021 and rapidly became the predominant strain^8,9^. Interestingly in our study, the initial few cases were of BA.1 (73.1%) while 26.8% were of BA.2 sub-lineage. This shift of sub-lineage in Omicron was also documented from Australia among 300 cases with travel history to South Africa^10^. The matter of concern is BA.2 sub-lineage, which has 303 additional unique mutations in comparison to BA.1. Of this mutational difference the most highlighting is the lack of spike protein deletion at amino acid 69/70 in BA.2, hence could not be detected by the SGTF(S-gene target failure) assay^10^.

We observed young adults and males were infected more in comparison to children and elderly population which could be because of more socialising habits and close connections than other mentioned groups. Our results suggest a large reduction in protection against COVID-19 infection with the Omicron variant as 87.8% population got reinfected after full primary vaccination thus implying increased breakthrough infections. This is in concordance with *in-vitro* studies that showed a significant reduction in neutralisation of the Omicron variant with convalescent sera or sera of fully vaccinated individual^11^. The data is too immature to analyse these breakthrough infections based on the type of the vaccine. However, it is worth mentioning that a population of 148.33 lakhs in Delhi had already completed its first dose of vaccination by December 24^th^ 2021 and two-third of them had been fully vaccinated with double doses^12^. Also the latest sero-survey study among Delhi population showed 89.5% seropositivity in the month of September-October 2021^6^. Amid increasing COVID-19 cases in the country, the Government on December 25^th^ had announced vaccination for the 15-18 year age group from January 3^rd^ 2022 and booster doses for healthcare and frontline workers from January 10^th^ 2022^13^.

Despite small numbers, we showed that individuals with a previous infection also had a clear increased risk of infection with Omicron compared with naïve individuals, suggesting that previous infection with another SARS-CoV-2 variant provides low levels of protection against Omicron infection. Our finding of reduced protection against reinfection with Omicron is in line with surveillance data from South Africa, showing increased risk of reinfections with the Omicron variant^14^.

Our findings strongly suggest that Omicron has a much higher rate of asymptomatic carriage resulting in high prevalence of asymptomatic infection, a likely major factor in the rapid dissemination of the variant locally and globally. Our results suggest a large decrease in protection from vaccine or natural immunity against COVID-19 infections caused by the Omicron variant. This emphasizes the urgent need for booster vaccination and will warrant implementing non-pharmaceutical interventions along with the installation of rapid detection strategies for asymptomatic carriage in high-risk transmission populations especially with those having comorbidities.

The majority of Omicron patients (60.9%) had no documented international travel history or contact hence undoubtedly acquired the infection locally, thus signifying the community spread and imposing further challenges in epidemic control. The study emphasized how the formation of local clusters from imported cases eventually led to community transmission. Now, we are on a war-footing to deal with the upcoming outbreak of the third wave of Covid-19. Parts of the world are approaching a transition or a new phase of the COVID-19 pandemic.

It is too soon to know the exact extent to which vaccination or previous infection will provide protection against reinfection with Omicron. The present study highlight several reasons for concern: (i) the rapid community transmission shortly after introduction of the first case in the National Capital with higher seropositivity rates (ii) High proportion of fully vaccinated individuals.

In our study we are able to document the initial few cases of an unusual rapid community spread by the Omicron variant in Delhi State. In addition our study provides an insight about clinical course in a population with high vaccination coverage and seropositivity against COVID-19 infection. Also, our data emphasized the role of near-real time genomic surveillance in the low-middle income countries as the Omicron community transmission surge continues, the virus evolves, and new variants with potentially altered fitness and bio medically relevant phenotypes might generate in near future.

The present data did not give a true magnitude of community transmission in Delhi State as positive cases from five districts had been studied. Therefore further studies incorporating wider population of national capital are warranted for better understanding the clinical course and epidemiology of this emerged variant in the community.

## Conclusion

To the best of our knowledge, this is the first study in which the early-stage representative data of local and community level transmission of infection from Delhi is investigated. Omicron infection was found to be associated with significantly increased breakthrough infections, decreased hospitalization rates, and less of symptomatic disease among individuals with high seropositivity against SARS CoV-2 infections. We observed a definite shift from Delta predominance to Omicron along with its community spread.

## Data Availability

All the sequences have been submitted to the GISAID database with hCoV-19/India/ILBS-WGSXXXX/2021

## Author Contributions

**Conceptualization:** E.G, A.A, S.K.S.; **Data curatio**n: R.A, R.G., P.G, V.S., A.B.; **Formal Analysis:** P.G., R.G.; **Funding acquisition:** E.G.,C.B.,A.A,S.K.S. **Experiments:** V.S., U.S.K,S.D.; **Project administration:** E.G.,C.B., S.K.S.; **Resources:** E.G., C.B., A.A, S.K.S.; **Supervision:** E.G., C.B., A.A, S.K.S.; **Visualization:** P.G.; **Writing – original draft:** R.G.,P.G., **Writing – review** & **editing: E.G**, R.G., P.G., R.A., S.K.S, A.A.

All authors have read and agreed to the published version of the manuscript.

## Data Availability Statement

All the sequences have been submitted to the GISAID database.

## Acknowledgments

We acknowledge the Government of NCT, Delhi for facilitating the Whole Genome Sequencing Laboratory for COVID-19 at our institute. We also extend our gratitude to National Liver Disease Biobank, ILBS for providing Next Generation Sequencing platforms.

## Research in context

### Evidence before this study

To identify existing evidence of community transmission, epidemiological and clinical characteristics of SARS-CoV-2 Variant of Concern (VoC) Omicron, we searched PubMed for peer-reviewed articles published between Nov 25, 2021, and Jan 04, 2022, using the keywords (“COVID-19” OR “SARS-CoV-2”) AND (“Omicron” OR “Variant”) AND (“Epidemiology” or “Community”) AND (“Infection” OR “Symptoms”). Our search was not restricted by language or the type of publication. We identified only one published PubMed article, which assessed the spread, epidemiology and clinical characteristics with severity of Omicron infections in Danish population. The study suggested that after its first appearance, Omicron variant, spread rapidly in Denmark, a European country with unique population characteristics such as high testing capacity, high vaccination coverage and limited natural immunity against SARS-CoV-2 infection. Spread was catalysed by super spreading events and warrants further epidemic control. Study showed the widespread community transmission from two travellers returning from South Africa. Most of the cases were symptomatic, while a few were hospitalized with no mortality.

### Added value of this study

To the best of our knowledge this is the first study from India to provide an early evidence of community transmission by newly identified Variant of Concern, Omicron. Initially restricted among international travellers and their contacts, Omicron spread within the community in a short span of time. The present study demonstrated the rapid community transmission among the population of New Delhi, a state with high seroprevalence and high rate of past infections against SARS-CoV-2. The study showed significantly increased breakthrough infections, decreased hospitalization rates, and lower rates of symptomatic infections among such cases.

### Implications of all the available evidence

Since the beginning of COVID-19, low and middle income countries like India have suffered a lot due to the upheaval caused by the pandemic. It has not just overburdened the healthcare system of the country but also exposed the gap and deficiencies in the medical facilities at community level. In the last couple of years COVID-19 has surmounted a huge toll of hospitalizations and deaths in the most densely populated states such as New Delhi. The numbers have been comparable to nothing seen since the Spanish flu. This fact coupled with astonishing high transmissibility of Omicron may push India on the verge of third wave and may even shatter the old records of daily cases.Keeping this in mind and the urgency of situation we were able to document the initial few cases of an unusual rapid community spread of Omicron variant in the national capital. In addition, our study will provide an insight about clinical course of the disease in a population with high vaccination coverage and seropositivity against COVID-19 infection laying the path for further progress.

Our data also emphasized the role of near-real time genomic surveillance in the low-middle income countries as the Omicron community transmission surge continues, the virus evolves, and new variants with potentially altered fitness and bio-medically relevant phenotypes might generate in near future.

